# Prospective prediction of clinical trial outcomes from public evidence

**DOI:** 10.64898/2026.11.17.26368242

**Authors:** Jie Xu, Wanheng Zhang, Ying Yuan, Chong Wu

**Affiliations:** Department of Biostatistics, The University of Texas MD Anderson Cancer Center, Houston, TX, USA; Department of Biostatistics and Data Science, School of Public Health, The University of Texas Health Science Center at Houston, Houston, TX, USA; Institute for Data Science in Oncology, The University of Texas MD Anderson Cancer Center, Houston, TX, USA

## Abstract

Predicting clinical trial outcomes requires interpreting biological and clinical evidence in the context of a specific intervention, population and study design. Here we test whether a general-purpose large language model (LLM) can predict trial outcomes from publicly available evidence. We introduce TrialDossier, which assembles standardized evidence dossiers, and TrialOutlook, which uses these dossiers with GPT-5 to estimate the probability that a trial meets its prespecified primary endpoint. Among 694 ongoing trials with results expected within six months of cohort assembly, 69 outcomes first disclosed after forecast recording had been adjudicated by 20 October 2026. TrialOutlook achieved an area under the receiver operating characteristic curve of 0.819 (95% confidence interval, 0.708–0.914) and an average precision of 0.860. TrialOutlook correctly identified 27 of 29 failures (93.1%) and incorrectly classified 17 of 40 successes as failures. We further recorded forecasts for 5,809 ongoing trials associated with US-listed companies and established a public benchmark linking archived evidence and fixed forecasts to adjudicated outcomes as they become available.

## Introduction

Clinical development requires decisions about investigational treatments based on incomplete biological and clinical evidence.^1–3^ We ask whether large language models (LLMs) can assemble and interpret publicly available biomedical evidence to predict individual trial outcomes before their public disclosure. Such prediction requires relating an intervention’s biological rationale and prior clinical findings to the trial’s population, comparator, endpoints and other design features. Reliable forecasts could help clinical development teams assess programme risk, identify where further evidence is needed and allocate resources.^4,5^

Human genetic support for drug targets is associated with clinical development success,^6–8^ and models integrating molecular, disease and trial information have predicted historical outcomes^9–11^ and prospectively forecast progression from phase 2 to phase 3.^12^ More recent systems based on LLMs have generated forecasts from trial protocols and biomedical evidence,^13,14^ while initiatives including CT Open^15^ and ClinicoBench^16^ provide frameworks for evaluating predictions as new outcomes emerge. These advances motivate a closer examination of how reliably LLM can forecast outcomes using the evidence available for ongoing trials, particularly when that evidence is incomplete, indirect or conflicting.

For an individual trial, relevant evidence is distributed across registries, publications, regulatory records and sponsor disclosures.^17,18^ A forecasting system must identify which findings apply to the intervention and clinical setting, reconcile differences in reporting and record where evidence is unavailable. Prospective evaluation requires forecasts recorded before public disclosure of outcomes.^19,20^ Performance estimates also depend on which trials enter the cohort and which yield adjudicable outcomes during follow-up. Trials with early readouts in a cohort selected for near-term results may not represent the broader population of ongoing trials. The practical relevance of forecasts also depends on the balance between missed failures and successful trials incorrectly predicted to fail.

Here we develop TrialDossier and TrialOutlook to assemble publicly available biomedical evidence and predict clinical trial outcomes. TrialDossier uses a trial identifier to compile a standardized dossier covering intervention biology, prior clinical findings, trial design, safety and operational characteristics. We assess the accuracy and coverage of dossier fields through manual review and comparisons with reference annotations, including those in ChEMBL^21^. TrialOutlook uses these dossiers with a general-purpose LLM to estimate the probability that a trial meets its prespecified primary endpoint. Among 69 eligible adjudicated readouts from an initial cohort of 694 trials, forecasts recorded before outcome disclosure discriminated between successes and failures. TrialOutlook identified 27 of the 29 failures while incorrectly classifying 17 of 40 successful trials as failures. To extend prospective evaluation beyond trials selected for near-term readouts, we recorded forecasts for a broader cohort of 5,809 ongoing trials conducted or sponsored by US-listed companies. We will report prospective forecast performance through the TrialOutlook website (https://trialoutlook.com/) that links evidence dossiers and recorded forecasts to adjudicated outcomes as results accumulate.

## Results

### Development and evaluation of an AI system for clinical trial outcome prediction

We first developed TrialDossier, a two-stage AI agent that uses a ClinicalTrials.gov identifier (NCT ID) to assemble a standardized evidence dossier for clinical trial outcome prediction (Fig. 1a; Methods). In stage 1, TrialDossier retrieves and extracts evidence from ClinicalTrials.gov,^17^ PubMed,^22^ Open Targets,^23^ openFDA,^24^ prespecified web sources and company disclosures, organizing it into 28 fields. Stage 2 standardizes or condenses eight fields and retains the remaining 20 without semantic rewriting. The resulting dossier covers intervention biology, prior clinical findings, trial design, safety and operational characteristics. Information that cannot be retrieved is explicitly recorded as missing.

**Fig. 1.**
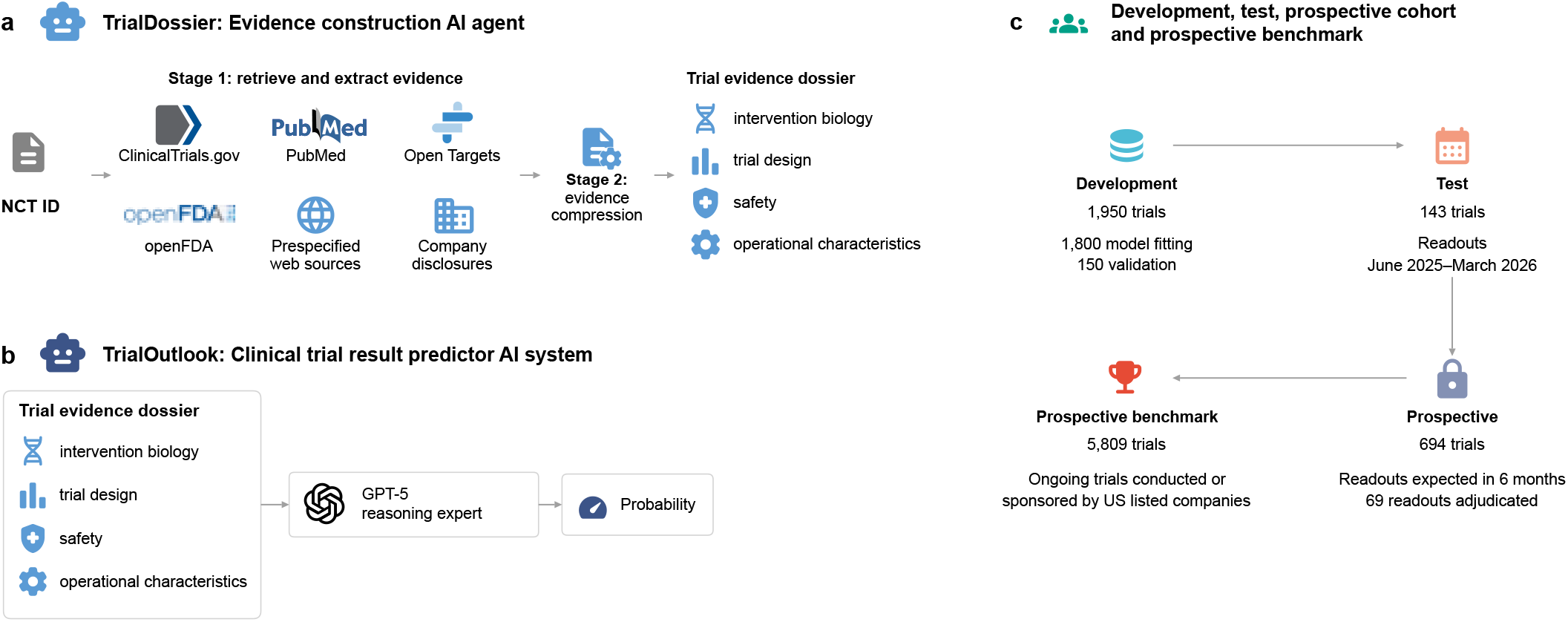
Overview of TrialDossier, TrialOutlook and the study cohorts. **a**, TrialDossier takes a ClinicalTrials.gov identifier (NCT ID), retrieves and extracts evidence from ClinicalTrials.gov, PubMed, Open Targets, openFDA, prespecified web sources and company disclosures (stage 1) and condenses it into a trial evidence dossier covering intervention biology, trial design, safety and operational characteristics (stage 2). **b**, TrialOutlook supplies the dossier to a GPT-5 reasoning expert, which returns the probability that the trial meets its prespecified primary endpoint. **c**, Study cohorts: the development cohort (1,950 trials; 1,800 for model fitting and 150 for validation), the retrospective test cohort (143 trials with readouts between June 2025 and March 2026), the initial prospective cohort (694 trials with readouts expected within six months, of which 69 were adjudicated) and the prospective benchmark (5,809 ongoing trials conducted or sponsored by US-listed companies).

TrialOutlook, our clinical trial outcome prediction system, uses these dossiers to estimate the probability that an individual trial meets its prespecified primary endpoint through a reasoning expert (Fig. 1b; Methods). Specifically, the reasoning expert uses GPT-5,^25^ a general-purpose LLM, prompted to update a phase specific base rate using the dossier evidence (Methods).

We also evaluate an embedding expert as an exploratory alternative. The embedding expert uses four text inputs, including the reasoning expert’s assessments of direct human evidence and of target and mechanism validation, functional summaries of target genes, and disease descriptions. A text embedding model^26^ converts these inputs into numerical vectors, and then TabPFN-3, a foundation model for tabular data,^27,28^ predicts clinical trial outcomes from these representations using labelled historical trials as reference examples (Methods).

We used four cohorts for model development, retrospective testing, and prospective evaluation (Fig. 1c; Methods). Outcome classification followed prespecified criteria appropriate to the source data. The development cohort comprised 1,950 trials with known outcomes (975 classified as successful and 975 as unsuccessful), including 1,800 fitting trials and 150 validation trials. Following model selection, the 1,800 fitting trials served as the reference set for TabPFN-3. To assess performance on trials excluded from model development, we used a separate retrospective test cohort of 143 trials with readouts first disclosed between June 2025 and March 2026, including 81 classified as successful and 62 as unsuccessful.

For an initial prospective assessment, we assembled 694 ongoing trials on 5 July 2026 with expected readouts within the following six months. Trials were identified through BioMedTracker, filings with the US Securities and Exchange Commission and company announcements; evidence was collected between 5 and 9 July 2026. Prospective comparisons included only adjudicated outcomes disclosed after the forecasts had been recorded (Methods). Early follow-up raised concern that successful trials could be overrepresented among the available readouts. To extend evaluation beyond trials selected for anticipated readouts, we assembled a broader prospective cohort of 5,809 ongoing trials in October 2026, comprising all eligible ongoing trials identified across US-listed companies at cohort assembly. We recorded forecasts before outcome disclosure and continue to evaluate performance as results become available.

### Accuracy, coverage and efficiency of TrialDossier

We compared TrialDossier with manual curation in a simple random sample of 20 trials from the prospective benchmark cohort of 5,809 trials (Methods). A PhD student in biostatistics, blinded to TrialDossier’s outputs, completed the same dossier schema using web searches, including Google Search and Gemini summaries. We restricted our comparison to the molecular target, target gene and mechanism of action (MOA), which require biological interpretation and restricted our comparisons to the prospective benchmark cohort to reduce the concern of knowledge leakage in LLM (Methods). For the 20 randomly selected trials, the human curator’s molecular target annotations were judged correct in 18 of 20 trials (90%), target gene annotations in 16 of 20 (80%) and MOA annotations in all 20 trials (100%). TrialDossier’s annotations were judged correct for all three fields in all 20 trials (100%) (Supplementary Tables 1 and 2). Furthermore, manual curation required a mean of 30.5 min per trial, corresponding to an estimated 353 h for all 694 trials. Using the July 2026 US average hourly earnings for private nonfarm employees ($37.62)^29^ as a wage benchmark, we estimated labour costs of $19.12 per trial. In comparison, TrialDossier’s LLM token cost was approximately $0.80 per trial (Fig. 2a).

**Fig. 2.**
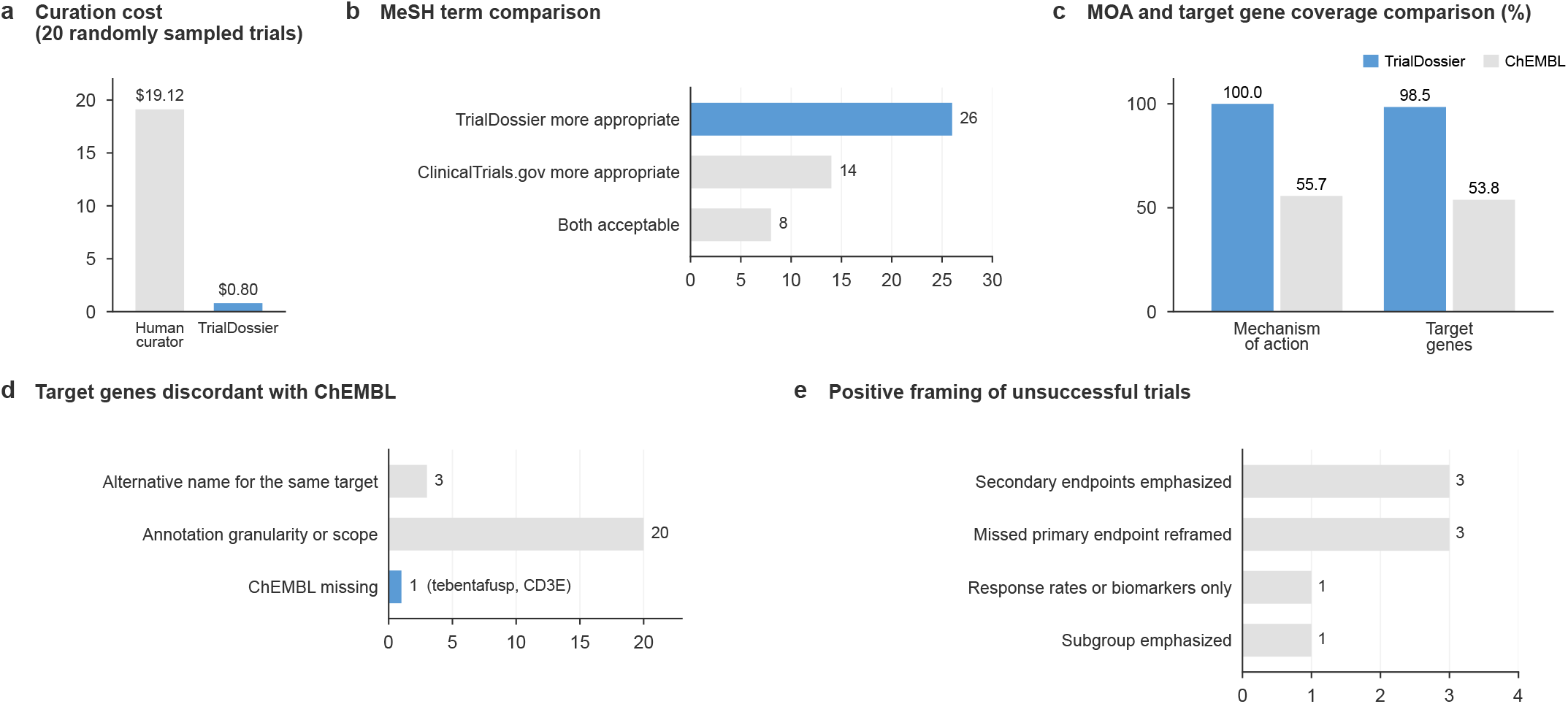
Evidence construction by TrialDossier. **a**, Estimated curation cost per trial of a human curator and of TrialDossier in 20 randomly sampled trials of the prospective benchmark. **b**, Adjudication of the 48 MeSH descriptors assigned by TrialDossier that differed from the single ClinicalTrials.gov descriptor, among 308 trials of the initial prospective cohort. **c**, Percentage of the 262 drug records of 200 randomly sampled trials of the prospective benchmark with MOA and target gene annotations from TrialDossier and ChEMBL. **d**, Groups of the 24 discordant target gene annotations among 131 trial–drug pairs matched in both sources; ChEMBL missing denotes a documented target recorded by TrialDossier and absent from ChEMBL (tebentafusp, CD3E). **e**, Dominant pattern of the 8 positively framed disclosures among 58 unsuccessful trials with readouts between June 2025 and March 2026. In b, d and e, bars show numbers of trials or trial–drug pairs.

To assess disease annotations across a larger sample, we compared the Medical Subject Headings (MeSH) terms^30^ assigned by TrialDossier with those in ClinicalTrials.gov. The comparison was restricted to the 308 trials for which the registry assigned a single MeSH term to the registered conditions in the initial prospective cohort. MeSH identifiers matched exactly in 260 trials (84.4%) (Supplementary Table 3). We manually reviewed the 48 discordant pairs to assess how well each term represented the registered conditions (Fig. 2b; Supplementary Table 4). In 24 pairs, one term represented a broader category and the other a more specific condition within that category. The more specific term was judged more appropriate in each case, favoring TrialDossier in 14 and ClinicalTrials.gov in 10. The remaining 24 pairs differed in the conditions or disease attributes represented, such as histology versus anatomic site, or involved errors in term or identifier assignment. Review favored TrialDossier in 12 cases and ClinicalTrials.gov in four; both annotations were acceptable in the remaining eight.

We further assessed the coverage and concordance of drug annotations by comparing TrialDossier with ChEMBL^21^. In a random sample of 200 trials from the prospective benchmark cohort, all 262 drug records named a drug intervention, representing 214 unique drugs. TrialDossier provided MOA descriptions for all 262 records (100%) and target gene annotations for 258 (98.5%). ChEMBL provided the corresponding annotations for 146 (55.7%) and 141 (53.8%) records, respectively (Fig. 2c; Supplementary Table 5).

Among 131 trial–drug pairs representing 92 unique drugs that were matched to ChEMBL by exact preferred name and had a target annotation in TrialDossier and a curated mechanism in ChEMBL, target gene annotations agreed directly in 107 pairs (81.7%) (Supplementary Table 6). Discordances were assessed using Claude Opus 5.5 and then reviewed manually against prescribing information and primary literature (Supplementary Table 7). No contradictory mechanisms were identified. Of the 24 discordant pairs, 3 reflected alternative names for the same target, and 20 reflected differences in annotation granularity or scope rather than conflicting mechanisms. These included annotations of entire protein families or complexes instead of specific drug-binding members, and differences in whether targets of antibody–drug conjugate payloads were included. For the remaining pair (tebentafusp), TrialDossier additionally annotated CD3 (CD3E), which is bound by the drug’s anti-CD3 domain on T cells (Fig. 2d; Supplementary Note 1).

We classified trial readouts using prespecified criteria applied to public disclosures (Methods). Success required evidence that the prespecified primary endpoint had been met, either through an explicit statement in the outcome disclosure or, when no such statement was available, by comparing the reported results with the primary endpoint registered on ClinicalTrials.gov. Failure required evidence of a missed primary endpoint or trial discontinuation without reported attainment of the primary endpoint. Outcomes remained unresolved when the evidence supported neither classification. We also assessed disclosure framing^31–33^ using a prespecified codebook, with framing judged by whether the dominant message of the disclosure (title, subtitle and conclusion) was favorable (Methods). Among 58 trials classified as unsuccessful with readouts between June 2025 and March 2026, the corresponding disclosures were classified as positively framed in 8 cases (13.8%), negatively framed in 48 (82.8%), and neutral in two (3.4%) (Fig. 2e; Supplementary Table 8). The positively framed disclosures emphasized secondary endpoints (3 trials), reframed a missed primary endpoint (3), or emphasized subgroup results (1) or response rates or biomarkers (1). For example, the sponsor of a phase 2b trial of zelicapavir in high risk adults with respiratory syncytial virus infection (NCT05568706) described the findings as “positive topline results” in the press release headline and highlighted a subgroup benefit, while disclosing the absence of a treatment effect on the primary endpoint only in the body of the release (Supplementary Note 2). These findings highlight the importance of adjudicating trial outcomes using prespecified criteria and explicit evidence.

### Development and retrospective evaluation of TrialOutlook

The reasoning expert used GPT-5 to estimate trial success probabilities from the TrialDossier dossier using a fixed prompt. To assess the contribution of the additional curated evidence, we compared the same GPT-5 model supplied with either the full dossier or a predefined trial profile from ClinicalTrials.gov in an additionally curated 150 trials from validation cohort (75 successes and 75 failures). Both conditions used the same outcome definition, prompt, and inference settings (Methods). Providing the full dossier increased ROC AUC by 0.034 (95% CI, 0.006–0.069) and AP (average precision) by 0.028 (95% CI, −0.001 to 0.059) relative to registry information alone (Fig. 3b; Supplementary Table 9).

**Fig. 3.**
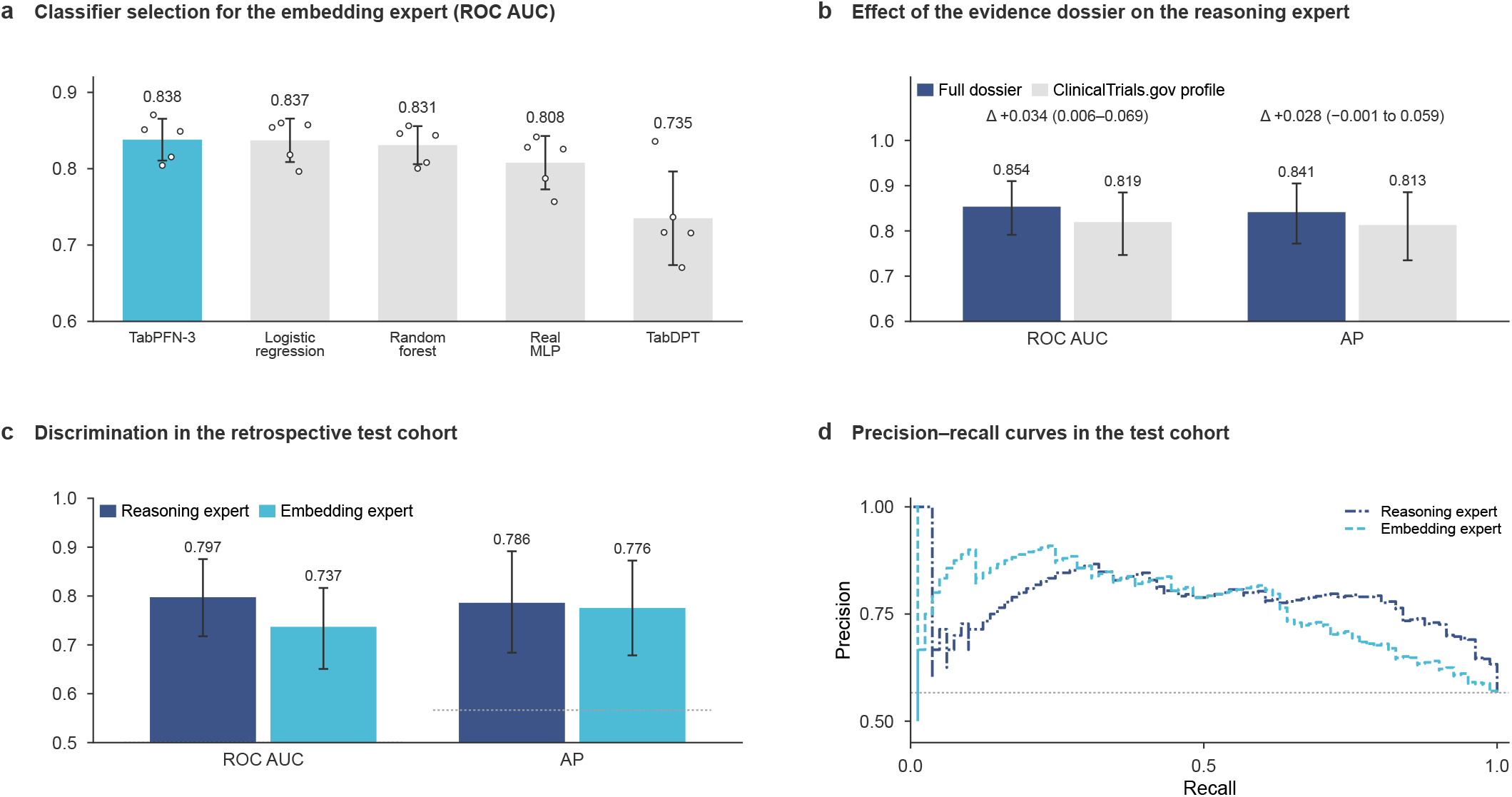
Development and retrospective evaluation of TrialOutlook. **a**, ROC AUC of five classifier families for the embedding expert in nested cross-validation grouped by primary investigational drug in the 1,800 fitting trials; bars and error bars show the mean and standard deviation across the five outer folds, and points show the individual folds. **b**, ROC AUC and AP of the GPT-5 reasoning expert given the full dossier or the ClinicalTrials.gov profile in the 150 validation trials, with the paired differences and their 95% CIs. **c**, ROC AUC and AP of the reasoning and embedding experts in the 143 retrospective test trials. **d**, Precision–recall curves of the two experts in the test cohort. Error bars in b and c show 95% bootstrap CIs; dotted lines in c and d mark the proportion of successful trials.

To extend the use of GPT-5 beyond direct probability estimation, we took its output assessments as inputs to an embedding expert. We developed this expert using the 1,800 fitting trials of the development cohort (Methods). The embedding expert encodes four text inputs, including the GPT-5 reasoning expert’s assessments of direct human evidence and of target and mechanism validation, functional summaries of target genes retrieved through MyGene.info,^34^ and disease descriptions associated with MeSH terms. This design was adapted from PRIORITI, which combines embeddings of biological rationales generated by an LLM with embeddings of gene summaries and disease definitions to forecast the success of target–indication pairs;^35^ here it is applied to individual trials. We compared five candidate classifiers using nested cross-validation grouped by primary investigational drug.^36^ TabPFN-3 achieved the highest area under the receiver operating characteristic curve (ROC AUC) (0.838) in the final cross-validation comparison across the 1,800 fitting trials and was selected for subsequent analyses (Fig. 3a; Supplementary Table 10). After classifier selection, the 1,800 fitting trials served as labeled reference examples for TabPFN-3.

We evaluated both experts retrospectively in 143 trials, comprising 81 successful and 62 unsuccessful trials, with results first disclosed between June 2025 and March 2026. These disclosures occurred after GPT-5’s reported knowledge cutoff of September 30, 2024.

We assessed discrimination using ROC AUC^37^ and the prioritization of successful trials using AP, with success as the positive class. The reasoning and embedding experts achieved ROC AUCs of 0.797 (95% CI, 0.718–0.876) and 0.737 (95% CI, 0.651–0.816), respectively, with corresponding AP values of 0.786 (95% CI, 0.684–0.892) and 0.776 (95% CI, 0.678–0.872) (Fig. 3c,d; Supplementary Tables 11 and 12).

We assessed the accuracy of the individual experts’ probability estimates using the Brier score and summarized calibration using expected calibration error (ECE).^38,39^ Brier scores were 0.189 (95% CI, 0.164–0.214) for the reasoning expert and 0.211 (95% CI, 0.176–0.249) for the embedding expert. Corresponding ECE values were 0.134 (95% CI, 0.104–0.213) and 0.107 (95% CI, 0.079–0.197), respectively (Supplementary Table 11). Although the embedding expert incorporated the reasoning expert’s assessments alongside gene and disease information, it achieved a lower ROC AUC than the reasoning expert. We therefore focused mainly on the reasoning expert in the prospective evaluation.

### Prospective evaluation of clinical trial forecasts

The initial prospective cohort comprised 694 ongoing trials identified through BioMedTracker, filings with the US Securities and Exchange Commission, and company announcements, with readouts expected within six months of cohort assembly. Forecasts from the reasoning expert were recorded on 29 July 2026. Only outcomes first disclosed after this date were included in the prospective evaluation, ensuring that all forecasts preceded their corresponding outcome disclosures (Fig. 4a). By 20 October 2026, results had been reported for 90 trials after the cutoff. We manually adjudicated these trials against prespecified outcome criteria, classifying 40 as successful and 29 as unsuccessful, while 21 remained unresolved (Supplementary Table 13).

**Fig. 4.**
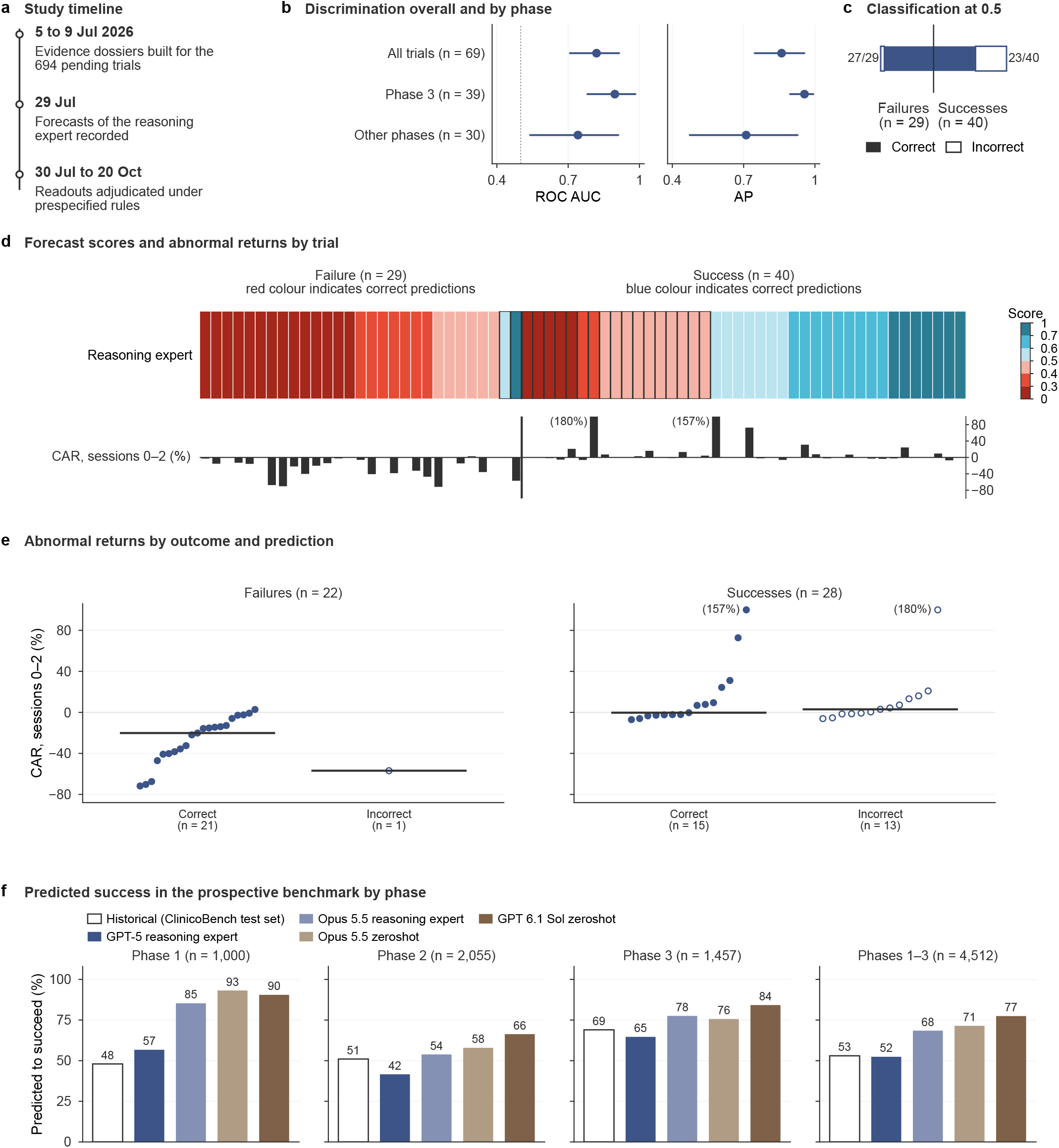
Prospective evaluation of TrialOutlook. **a**, Timeline of the initial prospective cohort. **b**, ROC AUC and AP of the reasoning expert in the 69 adjudicated trials (40 successes and 29 failures), overall and by development phase (exploratory), with 95% bootstrap CIs; the dotted line marks a ROC AUC of 0.5. **c**, Classification at the prespecified threshold of 0.5 by adjudicated outcome. **d**, Forecast scores of the reasoning expert for the 69 trials, ordered by score within each outcome (top; outlined cells mark incorrect predictions), and cumulative abnormal returns (CARs) over trading sessions 0–2 for the 50 eligible announcements (bottom). **e**, CARs of the 22 unsuccessful and 28 successful trials with eligible announcements by whether the prediction of the reasoning expert was correct (filled) or incorrect (open); horizontal lines show medians. In d and e, values beyond 100% are truncated, with the observed value in parentheses. **f**, Percentage of benchmark trials registered as phase 1, 2 or 3, and of the three phases combined, predicted to succeed (probability of at least 0.5) by the GPT-5 and Claude Opus 5.5 reasoning experts and the zero-shot forecasts of Claude Opus 5.5 and GPT-6.1 Sol, with the historical success rates of the ClinicoBench test set^16^ as reference.

Across the 69 adjudicated trials, the reasoning expert achieved a ROC AUC of 0.819 (95% CI, 0.708–0.914) and an AP of 0.860 (95% CI, 0.747–0.955) (Fig. 4b). The embedding expert achieved a ROC AUC of 0.813 (95% CI, 0.694–0.907) and an AP of 0.855 (95% CI, 0.741–0.938) (Supplementary Fig. 2; Supplementary Table 14). In an exploratory analysis, the reasoning expert had ROC AUC point estimates of 0.896 in phase 3 and 0.741 in other phases (Fig. 4b).

At the prespecified success probability threshold of 0.5, the reasoning expert correctly identified 27 of 29 failures (93.1%), while it incorrectly classified 17 successful trials as failures (Fig. 4c). Success predictions from the reasoning expert were correct in 23 of 25 cases (92.0%), although these captured only 57.5% of the 40 observed successes. With registry information alone, the same GPT-5 model and prompt achieved a ROC AUC of 0.796 (95% CI, 0.675–0.897), a difference of 0.023 (95% CI, −0.023 to 0.075) from the full dossier, but correctly classified 42 of 69 trials (60.9%) compared with 50 of 69 (72.5%) with the dossier (difference, 11.6%; 95% CI, 2.9–20.3; Supplementary Fig. 1).

We examined share price responses to outcome disclosures and whether they differed between trials whose outcomes the reasoning expert had predicted correctly or incorrectly.^40,41^ Of the 69 adjudicated prospective trials, 50 contributed eligible announcements (22 unsuccessful and 28 successful), after excluding trials with no listed equity exposure (n = 6), suspended trading (n = 1), trial status updated by the company without a clear news announcement (n = 2), multiple eligible trials disclosed in one release (n = 4), or other eligible announcements by the same company on the same event day (n = 6) (Supplementary Table 15). For each announcement, we estimated cumulative abnormal returns (CAR) over the first three trading sessions able to reflect the disclosure, using an XBI adjusted single factor model fitted to pre-announcement data. Event dates corresponded to the first disclosure supporting the adjudicated outcome (Methods). Negative CARs followed 21 of the 22 unsuccessful and 13 of the 28 successful trial announcements (Fig. 4d; Supplementary Table 16).

The reasoning expert correctly predicted failure for 21 of 22 unsuccessful trials with eligible announcements (median CAR, −20.2%), and the single unsuccessful trial predicted to succeed (NCT07108153) was followed by a CAR of −56.9%. Among successful trials, median CAR was −0.3% for the 15 correctly predicted to succeed by the reasoning expert and 3.0% for the 13 incorrectly predicted to fail (Fig. 4e). One illustrative example was MAXPIRe (NCT06132256), a phase 2 trial evaluating the CSF-1R antibody axatilimab in idiopathic pulmonary fibrosis. The reasoning expert assigned a success probability of 0.22. Although acknowledging axatilimab’s clinical activity in chronic graft-versus-host disease, the expert judged its relevance to pulmonary fibrosis to be limited, citing the historically high failure rate of phase 2 trials in this indication and the variability of the 26-week home spirometry endpoint (Supplementary Note 3). The trial subsequently failed to meet its primary endpoint, leading the sponsor to discontinue development in this indication. The failure announcement was associated with a CAR of −15.7%. As a sensitivity analysis, we repeated the analysis using all 62 of the 69 adjudicated prospective trials for which a CAR could be estimated, and the results were similar (Supplementary Note 4).

To evaluate performance beyond trials selected for anticipated readouts, we assembled a prospective cohort of 5,809 eligible ongoing trials across US-listed companies in October 2026, including trials without an announced readout date, and recorded forecasts before outcome disclosure (Methods). Of these trials, 1000 were phase 1, 2055 were phase 2 and 1457 were phase 3; the remaining 1297 comprised 730 phase 1/2, 168 phase 2/3, 65 early phase 1 and 334 phase 4 trials. For comparison, Chen et al.^16^ reported primary endpoint attainment rates in a retrospective cohort of trials initiated in 2021 or later were 48%, 51% and 69% for phase 1, phase 2 and phase 3 trials, respectively (Fig. 4f). To assess whether AI agents could predict clinical trial outcomes, we evaluated Claude Code with Claude Opus 5.5 as an additional reasoning expert using the same prompts and evidence dossiers as the GPT-5 reasoning expert. We also evaluated Claude Code with Claude Opus 5.5 and Codex with GPT-6.1 Sol as zero-shot forecasters on the same prospective cohort (Methods; Fig. 4f). At the 0.5 threshold, the GPT-5 reasoning expert predicted success for 52.4% of phase 1–3 trials, close to the historical rate of 53% for phases 1–3 combined, whereas the Claude Opus 5.5 reasoning expert predicted success for 68.5% and the zero-shot forecasts of Claude Opus 5.5 and GPT-6.1 Sol for 71.5% and 77.5%, respectively (Fig. 4f).

This live prospective benchmark that compares clinical trial forecasts as outcomes become available is at https://trialoutlook.com/. The benchmark preserves evidence dossiers and forecasts recorded before outcome disclosure, linking them to manually adjudicated outcomes and supporting sources. Original forecasts remain fixed as outcomes accumulate. Monthly updates compare our methods and external forecasts submitted before disclosure on shared eligible trials, summarizing discrimination, prioritization, classification errors, and, for probability forecasts, accuracy and calibration. Furthermore, SHA-256 hashes of the four forecast files are provided in this manuscript to enable verification of their integrity and detection of any subsequent modifications (Methods).

## Discussion

Our prospective evaluation shows that a general-purpose LLM supplied with publicly available biomedical evidence can discriminate subsequent clinical trial successes from failures. TrialDossier and TrialOutlook link forecasts recorded before outcome disclosure to standardized evidence dossiers and subsequently adjudicated outcomes, allowing both predictions and their supporting evidence to be examined. Differences in prediction targets, trial populations and information access limit direct comparisons with earlier evaluations of LLM forecasting.^13–16^

TrialDossier provides a standardized record of the evidence supplied to the forecaster, allowing input quality to be assessed alongside predictive performance. The full dossier improved discrimination relative to registry information alone in the historical validation cohort. A further comparison using archived inputs from the prospective cohort also showed higher classification accuracy at the prespecified threshold, although the estimated AUC gain remained uncertain.

TrialOutlook estimates the probability that a clinical trial meets its prespecified primary endpoint, an outcome that can be adjudicated at readout when sufficient results are available. Phase transition and regulatory approval concern broader development programmes that also depend on commercial priorities, development strategy and regulatory requirements.^1,2^ Thus, estimating phase transition probabilities or the probability of technical and regulatory success (PTRS)^10^ would require distinct prediction targets and evaluation. Also, TrialOutlook does not quantify the magnitude or clinical importance of treatment benefit, or its balance against harms. We leave these important topics to future research.

Interpreting these forecasts requires consistent outcome adjudication. Some sponsor disclosures framed unsuccessful trials positively,^31–33^ underscoring the need to apply prespecified success criteria to the reported results. Our adjudication rules also classified terminated trials as unsuccessful, although termination does not necessarily establish endpoint failure. Excluding the three terminated trials left 66 trials in the initial prospective benchmark, including 26 failures. ROC AUC and AP were 0.836 and 0.879, respectively, compared with 0.819 and 0.860 in the full 69 trial evaluation. The reasoning expert identified 25 of 26 failures after exclusion, compared with 27 of 29 in the full cohort. These results indicate that the observed discrimination was not driven by the inclusion of the three terminated trials.

The classification errors illustrate why discrimination must be interpreted alongside the consequences of individual predictions. At the prespecified success probability threshold of 0.5, the reasoning expert identified 27 of the 29 observed failures but also classified 17 of 40 successful trials as failures. In other words, 27 of the 44 failure predictions were correct (61.4%). A threshold suitable for identifying trials for additional scientific review may be inappropriate for decisions about continuing a development program. Using these probability estimates requires assessment of calibration and decision thresholds that reflect the consequences of missed failures and false warnings in the intended setting.^42,43^

In the exploratory market analysis, correctly predicted failures were accompanied by substantial negative abnormal returns, whereas successful trials elicited more variable and often modest responses,^41^ with no clear separation by forecast correctness. This pattern suggests that identifying likely clinical failures may have greater relevance to anticipating adverse market reactions. However, market responses also reflect investor expectations, commercial implications, and concurrent news. These observations characterize the market consequences of trial readouts but do not establish whether the forecasts provide information beyond investor expectations or could generate investment returns, which we leave to future research.

The initial prospective benchmark was limited to 69 adjudicable outcomes from trials selected for readouts expected within six months. A further 21 disclosures made after the forecasts remained unresolved. Disclosure timing and completeness may differ between successful and unsuccessful trials, affecting which outcomes contribute to performance estimates. Concern that available readouts might overrepresent successful trials motivated the broader cohort of 5,809 ongoing trials, which includes trials without an anticipated readout date. We expect this broader cohort will provide a less biased evaluation of whether general-purpose LLMs can predict clinical trial outcomes. We will update our prospective results with new readouts during revision.

As LLMs are updated, published trial outcomes may enter their training data, making performance on historical trials difficult to interpret when prior exposure to the results cannot be excluded.^15,20,44^ A dossier assembled before disclosure does not by itself resolve this problem if it is supplied to a newer model after the outcome is public. The benchmark therefore preserves both the evidence supplied and the forecast recorded before disclosure. This record allows the original predictions to be assessed regardless of what later models learn, while the dossiers and adjudicated outcomes can support further model development.

To maintain prospective evaluation as models evolve, we will repeat evidence collection annually for all benchmark trials, retain each dossier version and previous forecast, and generate new forecasts only for trials whose outcomes remain undisclosed. Comparing a fixed model version across successive dossiers will assess changes associated with updated evidence. Comparing model versions using the same dossier will assess changes associated with model updates, including changes in their underlying knowledge. All forecasts being compared will be recorded for the same trials before outcome disclosure, with matched access to external sources. Performance will then be evaluated as outcomes are adjudicated, with monthly updates to the public benchmark enabling continued assessment by the research community.^15,20^

In summary, our results demonstrate prospective discrimination of trial outcomes using public biomedical evidence. The benchmark will test whether this performance generalizes and improves as models and evidence are updated. Studies with clinical development teams could assess whether access to these forecasts improves assessment of programme risk and decisions about further evidence collection.

## METHODS

Methods are available in the **Methods** section.

## ACKNOWLEDGEMENTS

Research reported in this work was supported by National Cancer Institute under Award Numbers R01CA263494, U01CA293883, and P30CA016672; and Sabin Family Fellowship. The content is solely the responsibility of the authors and does not necessarily represent the official views of the National Institutes of Health.

## AUTHOR CONTRIBUTIONS

C.W. conceived and designed the study. J.X. and C.W. developed method. J.X. performed the analyses and wrote the initial version of the manuscript. W.Z. provided critical feedback and contributed to the analyses. All authors proofread the manuscript.

## Methods

### TrialDossier: evidence construction

TrialDossier used a two-stage workflow to assemble and standardize evidence for individual clinical trials. For the development, validation and retrospective test cohorts, it used Claude Sonnet 4.6 for retrieval, cleaning, and extraction in stage 1 and GPT-5.1 for standardization in stage 2; for the initial prospective cohort, it used Claude Sonnet 4.6 at both stages; for the expanded prospective benchmark, it used Claude Opus 5.5 at both stages.

Stage 1 accepted a ClinicalTrials.gov identifier (NCT ID) and produced a record containing 28 fields (Supplementary Table 17). The registry module queried the ClinicalTrials.gov v2 REST API^17^ and assembled the trial identifier, title, conditions, interventions, locations, principal investigator and affiliation, sponsor, recruitment status, phase, primary and secondary outcome measures, posted results and serious adverse events, allocation, masking, brief summary, eligibility criteria, study dates, investigator trial history, and protocol amendment history. Within this module, the LLM cleaned and deduplicated the intervention list and selected the core drug.

Downstream retrieval modules ran in parallel. The literature module queried PubMed Entrez Utilities^22^ for prior phase 1, phase 2, and preclinical reports of the core intervention. The regulatory module queried the openFDA REST API^24^ for approved indications, contraindications, warnings, adverse reactions, boxed warnings, and approval status. The investigator module summarized the principal investigator’s registered trial history. The disease module assigned a disease subcategory from the registered conditions and mapped it to a Medical Subject Headings (MeSH) term.^30^ The sponsor module used web search to characterize sponsor type, primary focus, and founding team expertise. The structure module retrieved the drug structure from PubChem,^45^ and a separate web search identified the molecular target.

Target resolution proceeded sequentially. A mechanism of action (MOA) resolver identified the intervention’s pharmacological action and candidate targets. A gene resolver mapped targets to human gene symbols approved by the HUGO Gene Nomenclature Committee (HGNC).^46^ The genetics module then queried the Open Targets Platform GraphQL API^23^ for target– disease associations, tractability, known drugs, safety, expression, interactions, and dependency evidence.

Stage 2 converted the retrieved record into a standardized dossier (Supplementary Table 17). Twenty fields were copied without semantic rewriting; eight were condensed or standardized and returned in fixed JSON schemas.

The genetic evidence field retained Open Targets information relevant to the trial indication. The endpoint profile removed repetition from outcome measures, endpoint families, and assessment windows. The eligibility profile organized criteria into four categories (population, inclusion, allowed, and exclusion) with entries specifying category, criterion, and value. It retained thresholds relevant to study design, including age, body mass index, performance status, and recent event windows, while removing consent, contraception, and routine laboratory boilerplate. Approved indication text was reduced to disease names, biomarker or subgroup qualifiers, and age limits.

Adverse reactions and boxed warnings were derived together from openFDA safety text. Boxed risks were represented by short labels; adverse reactions were grouped as common, serious but not boxed, or leading to discontinuation or death. Related events were combined into categories without repeating terms across lists. Investigator history was shortened by selecting up to eight titles, according to relevance to the trial disease, from up to 24 prior registered trials. Founding team expertise was summarized for up to four people, with normalized roles and short descriptions of relevant experience; finance, legal, and commercial roles were excluded. This was the only stage 2 field permitted to use additional web searches. All other fields were restricted to evidence retrieved in stage 1. Unavailable information was retained as missing rather than inferred or imputed.

### Development cohort

Development candidates were assembled from two historical pools. The earlier development pool comprised 2,104 records from a manually annotated 2020–2024 subset of the CTO benchmark^47^ and 657 records from BioMedTracker readout events and terminated ClinicalTrials.gov trials. Trials in the CTO subset retained their original outcome labels. BioMedTracker events reporting positive topline results were classified as successes, whereas trials listed as terminated in ClinicalTrials.gov were classified as failures. Excluding a previous holdout of 150 trials left 2,611 candidates.

The second pool was assembled from ClinicalTrials.gov records of completed trials with a completion date in January 2020 or later, an industry sponsor, a publicly listed parent company, and a specified phase (11,852 trials), together with 3,584 terminated trials. Completed trials were labeled using Claude Opus 5 in Claude Code. Subsequent trials were identified programmatically, and every success label was verified through web search. Terminated trials were labeled as failures on the basis of registry status. We excluded trials without a clear outcome signal, trials in the retrospective test cohort, trials whose success labels could not be confirmed on repeat verification, trials enrolling healthy volunteers, and records without an intervention. This yielded 4,222 trials: 1,515 successes and 2,707 failures.

From the pooled candidates, the 900 successes and 900 failures with the most recent completion dates formed the fitting set of 1,800 trials. A separate validation set of 150 trials, comprising 75 successes and 75 failures, was sampled at random from the remaining candidates completed during 2020–2023 after excluding the retrospective test. The validation set contained 19, 19, 19, and 18 successes and 23, 15, 14, and 23 failures completed in 2020, 2021, 2022, and 2023, respectively. Together, the fitting and validation sets comprised 1,950 trials. Classifier comparison used the 1,800 fitting trials; the 150 validation trials were used for the comparison of the reasoning expert with and without the full evidence dossier.

### Retrospective test cohort

The retrospective test cohort comprised 143 trials with results first disclosed between June 2025 and March 2026: 81 successes and 62 failures. Candidate readout events were obtained from BioMedTracker. Events were adjudicated using the outcome rules described below.

### Initial prospective cohort

The initial prospective cohort was assembled from trials expected to report results within six months. The BioMedTracker calendar supplied 1,000 upcoming topline result events. Of these, 686 entries were mapped to ongoing ClinicalTrials.gov records, representing 525 unique trials; 496 trials were retained after dossier construction and exclusions. Searches of recent filings from more than 1,000 US listed sponsors identified 349 unique trials expected to report results during 2026. Of these, 210 were not already present in the BioMedTracker derived set and were added. Trials were excluded if they were not ongoing at assembly or could not be mapped to a unique registry record. Matching NCT IDs against the development and retrospective test cohorts identified nine and three overlapping trials, respectively, which were removed, leaving 694 trials.

### Expanded prospective benchmark cohort

We queried the ClinicalTrials.gov v2 API for trials with an ongoing status: recruiting, not yet recruiting, active and not recruiting, or enrolling by invitation. The query returned 121,961 records. We retained trials whose lead sponsor or collaborator was a company listed on the New York Stock Exchange, Nasdaq, NYSE American, or Cboe, including listings through American depositary receipts and subsidiaries mapped to their listed parent (10,607 trials). Restriction to interventional studies left 8,686 trials; requiring a specified phase, including early phase 1, left 7,418. Excluding trials enrolling healthy volunteers left 6,855.

We checked by web search the 1,736 trials whose primary completion date had passed or that had posted results. Removing 872 trials with a disclosed final result or an interim stop left 5,983. We then excluded 86 trials whose dossiers listed no intervention, 20 trials present in the development cohort, 25 in the retrospective test cohort, and 43 from the initial prospective cohort whose readouts had been disclosed before forecast recording. The final benchmark contained 5,809 trials.

### Manual curation and cost benchmark

A PhD student in biostatistics completed the same dossier schema for a simple random sample of 20 trials from the expanded benchmark of 5,809 trials. The curator received only the NCT IDs, was masked to TrialDossier outputs, and could consult ClinicalTrials.gov, the open web, and publicly accessible databases, including Google Search and Gemini summaries. Curation time was recorded for each trial. TrialDossier annotations were taken from the corresponding benchmark dossiers. Registry derived variables were not compared because registry records can change after posting and the website and API do not present identical content. The annotation comparison was restricted to molecular targets, target genes, and MOA.

A molecular target annotation was considered correct if it identified the primary pharmacological target of the core intervention. Target gene annotations were assessed against HGNC human gene symbols; protein aliases were not accepted as gene symbol matches. For interventions without a human gene target, a not applicable value, a blank value, or a non-gene entry was accepted. An applicable annotation was considered incorrect if any listed entry was wrong or no target was supplied. Incorrect entries included the drug itself, the gene encoding a protein drug or a symbol not approved by HGNC. For bispecific antibodies and T cell engagers, both bound antigens were accepted. For gene therapies and genetically modified cell products, the transgene and its product were accepted. An MOA annotation was correct if it identified the therapeutic target or modality without factual error and incomplete if it provided only a general description identifying neither.

Manual effort for the initial cohort was estimated by multiplying the mean curation time in the 20 trial sample by 694. Labor cost was calculated from elapsed hours using the July 2026 US average hourly earnings for private nonfarm employees of $37.62.^29^ Automated expenditure was expressed as LLM token cost per trial and extrapolated to the same cohort.

### MeSH audit

MeSH assignments were compared with the ClinicalTrials.gov MeSH module in the 308 trials of the initial prospective cohort for which the registry assigned a single term to the registered conditions. Assignments were concordant when their MeSH identifiers matched exactly. One author reviewed discordant pairs against the registered conditions and eligibility criteria and classified each pair as favoring TrialDossier, favoring ClinicalTrials.gov, or supporting both terms. A random sample of approximately 10% of concordant pairs was reviewed using the same procedure to identify clinically inappropriate assignments despite matching identifiers (Supplementary Tables 3 and 4).

### Drug mechanism and target gene audit

The drug annotation audit used a random sample of 200 trials from the expanded prospective benchmark. Trial identifiers were sorted in ascending order, and two consecutive samples of 100 were drawn without replacement using one random number generator with seed 20261028. Each core intervention in a sampled trial constituted one drug record. ChEMBL release 37 annotations were retrieved through its web services, including molecular targets and target genes.^21^ Coverage was calculated separately for MOA and target gene annotations as the proportion of eligible drug records with a valid annotation. ChEMBL matching for coverage used preferred names, synonyms, or validated searches. Direct concordance analysis was restricted to records matched by exact preferred name that had a target annotation in TrialDossier and a curated mechanism in ChEMBL (Supplementary Tables 5–7).

Gene annotations were split on delimiters and represented as HGNC symbols. Records were concordant if their normalized target names matched or, when names differed, their gene sets were identical; the latter were recorded as nomenclature or alias differences. Partial overlap, including subset relationships, was flagged as a potential difference in annotation scope. Records for which neither target names nor associated genes could be mapped reliably received the highest review priority.

Claude Opus 5.5 reviewed flagged records using both sources’ annotations, mechanism descriptions, and associated evidence. It assigned a predefined category with an evidence-based justification: alias or standard name differences; family or class enumeration; antibody– drug conjugate payload representation; perspective or granularity differences; modality representation; a documented target absent from ChEMBL; an absent or unknown ChEMBL mechanism despite supporting evidence; pharmacological direction or terminology differences; or genuine biological disagreement. The first five categories and pharmacological direction or terminology differences were treated as biologically reconcilable. Missing ChEMBL annotations were recorded as gaps, and genuine biological disagreement remained unresolved.

An author subsequently checked every model adjudicated record against the cited prescribing information or primary literature. Manual decisions superseded model classifications, and the final classification and rationale were retained. Annotation conventions affecting multiple records were recorded as systematic differences rather than independent errors. A random sample of approximately 10% of automatically concordant records underwent the same manual review.

### Sponsor disclosure framing

We obtained BioMedTracker events dated June 2025 to March 2026, of which 58 were labeled unsuccessful.

Disclosure framing was classified as positive, negative, or neutral or unclear based on the dominant message conveyed by the title, subtitle, and conclusion concerning the target trial, drug, and indication. Positive framing emphasized favorable efficacy or clinical findings; negative framing emphasized endpoint failure, lack of efficacy, safety concerns, or development discontinuation; and neutral or unclear framing included process updates, disclosures without outcome claims, or mixed messages. For positively framed disclosures, we additionally recorded whether primary endpoint attainment was explicitly reported and the basis for favorable framing, including secondary endpoints, subgroup findings, or favorable interpretations of missed primary endpoints. All classifications were subsequently manually reviewed by the authors against the source disclosures.

### Trial outcome adjudication

Trials were classified as successful or unsuccessful according to whether the reported results satisfied the prespecified criteria for the primary endpoints. Explicit statements of primary endpoint attainment or failure in trial readouts were used when consistent with the trial’s prespecified design. When such statements were absent or ambiguous, the authors compared the reported results with the primary endpoints documented in ClinicalTrials.gov. Trials terminated before completion were classified as unsuccessful, unless attainment of the prespecified primary endpoint had been reported.

For trials with multiple primary endpoints, adjudication followed the prespecified criteria for overall trial success. Trials requiring all co-primary endpoints to be met were classified as unsuccessful if any required endpoint failed, whereas trials permitting success on a subset of primary endpoints were evaluated according to their prespecified multiplicity-adjusted testing strategy. Trials were classified as unresolved when the available results or prespecified decision criteria were insufficient to determine whether the primary endpoints had been met, or when conflicting information could not be resolved.

### Reasoning expert

The reasoning expert used GPT-5 through Azure OpenAI Service. The model received the dossier fields listed in Supplementary Table 17. The GPT-5 reasoning expert had no web search or other external retrieval access during inference; its reported knowledge cutoff was September 30, 2024.

The model estimated the probability that the trial would meet its prespecified primary endpoint. Preprocessing recorded the trial phase and assigned clinical and therapeutic characteristics using predefined rules. Evidence was organized into eight domains covering clinical, preclinical, mechanistic, trial design, class precedent, population, sponsor, and evidence quality considerations.

Fixed prompts instructed the model to update a base rate using the supplied evidence (Supplementary Note 5). The base rate was fixed at 48% for phase 1 trials, whereas for other phases the model estimated a base rate interval from its own knowledge. Strong direct clinical evidence or disqualifying findings warranted larger probability updates, while indirect or uncertain evidence received less weight.

The model returned success probability intervals, supporting and opposing evidence, uncertainties, and a rationale. Each run’s forecast was the midpoint of its success probability interval. Four runs were initiated per trial, and their successful outputs were averaged to obtain the reasoning expert’s forecast.

On the expanded prospective benchmark, Claude Opus 5.5 was also run as a reasoning expert in Claude Code at extra-high thinking effort, with the prompts of the GPT-5 reasoning expert (Supplementary Note 5), the same dossier fields and no tool or web access.

### Contribution of curated evidence

The reasoning expert received either the full dossier or registry information from the 150 validation trials. Both used the same GPT-5 model, prompt, and inference settings. The registry condition supplied ten fields: NCT ID, phase, core intervention, disease, endpoint profile, randomization, blinding, eligibility profile, sponsor, and sponsor type. Endpoint and eligibility profiles retained their stage 2 condensed representations. Each condition was run four times per trial, and run probabilities were averaged. Paired differences in the area under the receiver operating characteristic curve (ROC AUC) and average precision (AP) were estimated from 2,000 bootstrap resamples stratified by outcome, using seed 2, with 95% percentile confidence intervals (Supplementary Table 9). In the 69 adjudicated prospective trials, the registry condition was evaluated post hoc. Registry forecasts were generated after the outcomes had been adjudicated, from the same ten fields of the dossiers used for the prospective forecasts, with four runs per trial, and were compared with the reasoning expert’s prospective forecasts. Differences in ROC AUC and in the proportion of trials correctly classified at the prespecified threshold of 0.5 were estimated with the same stratified bootstrap.

### Embedding expert

Each trial was represented by a vector of 1,792 dimensions. The first 768 dimensions encoded the reasoning expert’s assessments. In each of the four reasoning runs, the assessment of direct human evidence and the assessment of target and mechanism validation were joined into one text, which was embedded with Gemini embedding 001 at 768 dimensions using the classification task type. Each run vector was normalized to unit L2 norm, the vectors of runs with assessment text were averaged, and the mean was normalized again; a trial without assessment text in any run received a zero vector.

A further 512 dimensions encoded functional summaries retrieved through MyGene.info for HGNC symbols parsed from the target gene field. Another 512 dimensions encoded the scope note of the trial’s MeSH term, retrieved from the National Library of Medicine MeSH service. The MeSH term was mapped from the dossier’s disease field using the procedure of the TrialDossier disease module. If no term was available, the scope note was sought by disease name. If no scope note was available, the MeSH term or, failing that, the disease name was embedded. A gene symbol replaced its functional summary when MyGene.info returned no summary. A single space was embedded when no target gene or disease text was available. Target and disease embeddings were generated once per trial with Gemini embedding 001 at 512 dimensions and the classification task type, then normalized to unit L2 norm.

TabPFN-3^27,28^ classified the resulting vectors using the v3 classifier checkpoint in tabpfn 8.0.8. Settings were defaults except that limits on reference set size and the number of features were disabled with ignore_pretraining_limits. The embedding expert was applied to the retrospective test cohort and the initial prospective cohort, with random seed 42 for both. At inference, TabPFN-3 conditioned on labeled reference examples without gradient updates to the model parameters. The same 1,800 fitting trials were used as the reference set for every cohort.

### Classifier comparison

Five classifier families were compared using the same features in the 1,800 fitting trials: TabPFN-3, random forest,^48^ RealMLP (pytabkit 1.7.3),^49^ logistic regression, and TabDPT (tabdpt 1.2.0).^50^ Trials were grouped by primary investigational drug, defined as the first normalized noncomparator entry in the core intervention field. This produced 1,315 groups; ten trials without a drug name each formed a separate group. The outer cross-validation loop used five folds stratified by outcome and grouped by drug, with seed 42. No primary drug appeared in both the fitting and evaluation portions of an outer split.

Within each outer fitting split, a grouped inner loop with three folds selected hyperparameters by ROC AUC for random forest and logistic regression. Random forests used 500 trees, maximum features of either the square root of the feature count or 0.1 of the features, and minimum leaf sizes of 1 or 5. Logistic regression used standardized inputs and C values of 0.001, 0.01, 0.1, or 1. TabPFN-3, TabDPT, and RealMLP used default settings. Classifier performance was summarized by the mean and standard deviation of ROC AUC across the five outer folds; TabPFN-3 had the highest mean and was selected (Supplementary Table 10).

### Prospective evaluation

For each of the 694 initial prospective trials, Claude Opus 5.5 in Claude Code identified public announcements of primary or topline results or trial discontinuation. An author manually checked every provisional outcome against the linked source and finalized labels using the criteria for recent readouts.

By October 20, 2026, 155 trials had reported results or discontinuation and 539 had no final result. We excluded 65 trials with disclosures before 29 July 2026. Of the remaining 90, nine lacked sufficient evidence for either outcome, seven reported an endpoint that did not match the registered primary endpoint, and five reported interim data only. These 21 trials remained unresolved. The evaluation therefore included 69 adjudicated trials: 40 successes and 29 failures (Supplementary Table 13).

### Share price responses to disclosures

The announcement analysis began with the 69 adjudicated prospective trials. For each, we identified the listed company with the greatest exposure to the outcome and verified the date and time of the first public disclosure supporting the label. Event day 0 was the first trading session able to reflect the disclosure: the announcement date for releases before the open or during regular trading, and the next trading day for releases after the close or on a nontrading day. We excluded six trials without listed equity exposure, one with suspended trading, two whose trial status was updated by the company without a clear news announcement, four whose releases reported multiple eligible trials, and six with another eligible announcement by the same company on the same event day. The analysis contained 50 announcements: 22 unsuccessful and 28 successful trials (Supplementary Table 15).

Abnormal returns were estimated using a single factor market model with the SPDR S&P Biotech exchange traded fund (XBI) as the index. Ordinary least squares was fitted to daily simple returns calculated from adjusted closing prices over trading days −130 through −11 relative to day 0. Cumulative abnormal returns (CARs) were obtained by summing the differences between observed and model predicted daily returns over days 0–2 for the primary analysis and days 0–1 and 0–5 for alternative windows. For listings outside the United States, local closing prices were paired with the XBI close on the same calendar date. Announcements accompanied by other material company news during the event window, including financial results, financing, or results from other trials, were flagged; the analyses were repeated after excluding them. CARs were compared between trials whose outcome the reasoning expert had predicted correctly or incorrectly at the 0.5 threshold, separately for successful and unsuccessful trials. Differences in median CAR (correctly minus incorrectly predicted) were summarized with 95% percentile bootstrap intervals from 2,000 resamples drawn with replacement within each group. In a sensitivity analysis, the comparison was repeated in all 62 adjudicated prospective trials for which a CAR could be estimated, including the announcements excluded above (Supplementary Note 4).

### Continuous prospective benchmark

The prospective cohorts are maintained at https://trialoutlook.com/. Outcomes follow the criteria for recent readouts and are updated monthly. External submissions containing the NCT ID, predicted success probability, method name, and generation date. To enable verification of the integrity of the expanded prospective benchmark forecasts and detection of any subsequent modifications, SHA-256 hashes for each of the four forecast files—corresponding to the GPT-5 and Claude Opus 5.5 reasoning experts and the Claude Opus 5.5 and GPT-6.1 Sol zero-shot forecasts—are provided alongside the data and code. The SHA-256 hashes, shown in groups of eight characters, are as follows: 5b250840 825a7cea b4216c1d 59dd230c 7f3e63ba b94091fc 0d1f51d1 feeb3ab4 (GPT-5 reasoning expert); 454687c7 57b6c28b 7e34e64f 4ba77f79 65bfdcd0 89496e1e f9b6bcf8 58696cf2 (Claude Opus 5.5 reasoning expert); fe20ef40 2a9f529d d8698e7c 0073c28b 96de03fc d93fa83d 8261ab68 c9c20b61 (Claude Opus 5.5 zero-shot forecasts); and cd2bd2cd 8d1242e9 83b89675 7cfa6936 2e54d35a cf37edad 18cb9537 20b5e4ea (GPT-6.1 Sol zero-shot forecasts).

Submitted methods and TrialOutlook are compared on shared eligible trials using ROC AUC, AP, and classification errors. The Brier score and expected calibration error are evaluated for probability forecasts. Updates include uncertainty estimates, forecast coverage, and the numbers of adjudicated, unresolved, and pending trials. Original forecasts remain fixed as outcomes accumulate.

### Zero-shot forecasts

For each trial of the expanded prospective benchmark, GPT 6.1 Sol was run in the Codex command line interface at medium thinking effort, in a separate session per trial, with the NCT ID as the only input. The fixed prompt (Supplementary Note 6) allowed live web search. The model returned an integer success probability from 0 to 100 with up to three reasons. Claude Opus 5.5 was run in Claude Code at medium thinking effort under the same conditions, with the prompt given in Supplementary Note 6, and returned the same output.

## Statistical analysis

Success was the positive class. Discrimination was summarized by ROC AUC, calculated from the Mann–Whitney statistic with half credit for tied scores, and by AP. Probability accuracy was assessed with the Brier score and calibration with expected calibration error calculated using ten bins of equal width. Sensitivity, specificity, positive predictive value, and negative predictive value used the 0.5 cutoff. With success as the positive class, specificity denotes the proportion of unsuccessful trials classified as failures.

Ninety-five percent confidence intervals for ROC AUC, AP, the Brier score, and expected calibration error were percentile bootstrap intervals from 2,000 resamples of trials with replacement, using seed 42. Resamples containing only one outcome class were discarded. Intervals for single proportions were two sided Clopper–Pearson exact intervals. The comparison of evidence conditions used its separately specified stratified bootstrap with seed 2. No adjustment was made for multiple comparisons.

Exploratory phase analyses compared trials registered as phase 3 with those registered as phase 1, phase 1/2, phase 2, or phase 2/3. Phase specific outcome proportions retained the registry phase categories. Analyses used Python 3.12.4 with NumPy 1.26.4, SciPy 1.13.1, scikit-learn 1.4.2, pandas 2.2.2, statsmodels 0.14.2, and matplotlib 3.8.4.^51–56^

## Computing environment

GPT-5 was accessed through Azure OpenAI Service and Gemini embedding 001 through the Google Generative AI API. Classifier comparisons ran on the MD Anderson Seadragon cluster with an NVIDIA L40 GPU, tabpfn 8.0.8, tabdpt 1.2.0, pytabkit 1.7.3, scikit-learn 1.7.2, and torch 2.4.1. Embedding expert predictions were generated on a workstation with an NVIDIA GeForce RTX 4070 GPU using Python 3.11.18, tabpfn 8.0.8, torch 2.5.1 with CUDA 12.1, scikit-learn 1.7.2, NumPy 2.3.5, and pandas 2.3.3. Prompts and dossier fields are referenced in Supplementary Note 5 and Supplementary Table 17.

## Data availability

The cohort manifests, evidence dossiers, model outputs and forecasts, adjudicated outcomes with their sources, curation audit records and analysis outputs are provided for peer review in the accompanying Replication_codes_and_data.zip and will be publicly available at https://github.com/ChongWuLab under a Creative Commons Attribution-NonCommercial 4.0 International licence (CC BY-NC 4.0) upon publication. The prospective benchmark, with dossiers, forecasts and adjudicated outcomes updated as trials read out, is maintained at https://trialoutlook.com/. Trial records were obtained from ClinicalTrials.gov, and further evidence from PubMed, Open Targets, openFDA, PubChem, HGNC, MyGene.info and MeSH, which are publicly available. Data from third parties retain their original licences; ChEMBL annotations (release 37) are provided under CC BY-SA 3.0. BioMedTracker (Citeline) is available by subscription. Daily share prices were obtained from Yahoo Finance and checked against Nasdaq. Source Data are provided with this paper.

## Code availability

Code for TrialDossier, TrialOutlook and all analyses is provided for peer review in the accompanying Replication_codes_and_data.zip for review purpose and will be publicly available at https://github.com/ChongWuLab under CC BY-NC 4.0 upon publication. The script code/run_all_local.py regenerates all results, figures, Supplementary Tables and Source Data from the deposited model outputs without API access. Building dossiers and running the reasoning and embedding experts require the user’s own Anthropic, Azure OpenAI and Google API access. For users without Azure access, we also provide a standard OpenAI API implementation with identical prompts, which was not used in this study. Prompts of the reasoning expert are given in Supplementary Note 5. The TabPFN-3 classifier weights included in the repository are an unmodified copy distributed under the TABPFN-3 License v1.0 for noncommercial use.

## Figure legends

**Supplementary Fig. 1.**
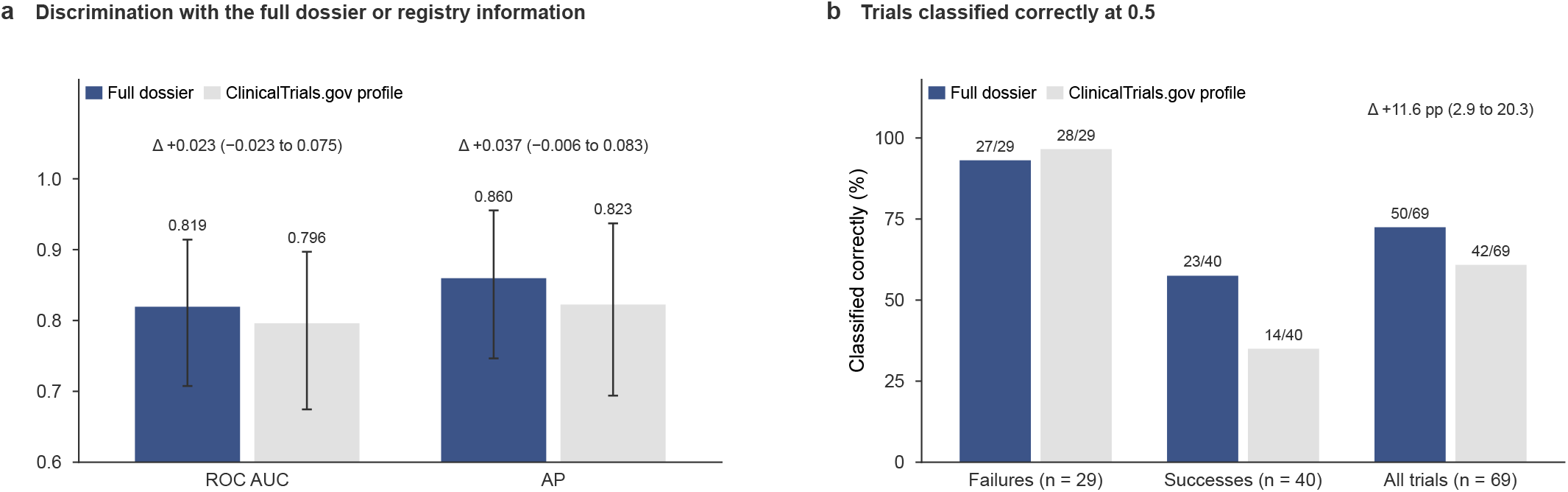
Full dossier versus registry information in the initial prospective cohort. **a**, ROC AUC and AP of the GPT-5 reasoning expert given the full dossier or the ClinicalTrials.gov profile in the 69 adjudicated trials, with 95% bootstrap CIs and the paired differences with their 95% CIs. **b**, Percentage of failures, successes and all trials classified correctly at the prespecified threshold of 0.5; numbers above the bars give the correct classifications, and the difference in overall accuracy is shown with its 95% CI.

**Supplementary Fig. 2.**
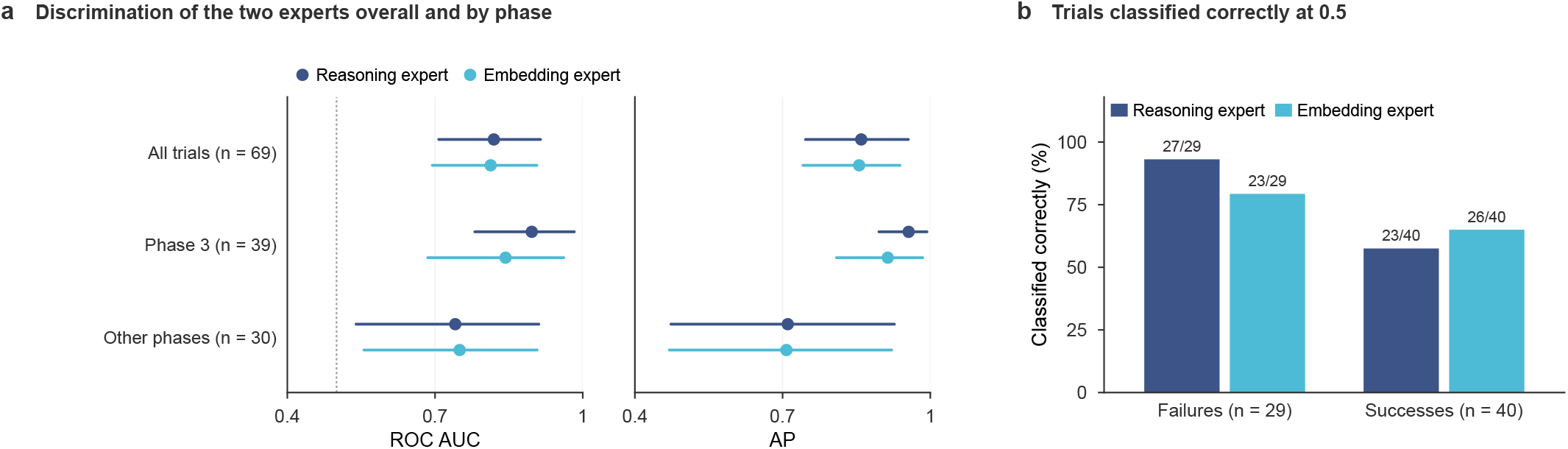
Prospective performance of the embedding expert. **a**, ROC AUC and AP of the reasoning and embedding experts in the 69 adjudicated trials, overall and by development phase (exploratory), with 95% bootstrap CIs; the dotted line marks a ROC AUC of 0.5. **b**, Percentage of failures and successes classified correctly at the prespecified threshold of 0.5; numbers above the bars give the correct classifications.

